# Identifying opportunities for improving the organ supply through race-stratified data

**DOI:** 10.1101/2021.10.24.21265203

**Authors:** David Goldberg, Darius Chyou, Brianna Doby, Raymond Lynch

## Abstract

Organ procurement in the US has received attention from government officials and policymakers the last two years, culminating in CMS releasing an updated Final Rule related to organ donation this year. This regulatory change revises how organ procurement organizations (OPOs), the federal contractors tasked with managing deceased donation, are evaluated and certified/de-certified. We used 2019 data and the CMS methodology to calculate race-stratified donation data among racial/ethnic minorities across the 57 OPOs. We found that the variability in donation rates across the 57 OPOs are greater among minority populations than non-Hispanic white potential donors. Among Tier 3 OPOs, there are: a) some with low donation rates across all racial/ethnic groups; b) some with low donation rates among only certain groups, and c) some where donation rates are lowest among non-Hispanic white patients. Among low-performing OPOs, these race/ethnicity-stratified data show that under-performance in certain areas is not due to the population demographics, and identifies areas for targeted interventions to increase donation and avoid decertification

## Introduction

Organ procurement in the US has received attention from policymakers, culminating with this year’s update to the Centers for Medicare and Medicaid Services (CMS) ‘Final Rule.’^1,2^ With this change, organ procurement organizations (OPOs), the 57 federal contractors tasked with managing deceased donation in zones known as donation service areas (DSAs), will be assigned to tiers based on the upper limit of the one-sided 95% confidence interval around their recovery and transplantation rates.^1,2^ At the end of each four-year cycle, Tier 3 OPOs will be decertified, but the policy’s use of confidence limits could render all OPOs as Tier 1 or 2 (one-sided 95% CI at or above median).^1,2^ Identifying underserved areas and populations is critical to increasing the organ supply and avoiding OPO decertification. We sought to explore OPO-level variability in procurement rates within racial/ethnic categories to identify high priority areas and groups for focused quality improvement.

## Methods

We conducted a retrospective cohort study to calculate OPO-specific ‘donation’ rates (donors recovered per 100 donation-consistent deaths) using CMS methods: a) donors (patient with ≥1 organ transplanted) using Organ Procurement and Transplantation Network data; and 2) potential donors using National Center for Health Statistics Multiple Cause of Death data.^1,3,4^ To avoid small sample size bias, DSAs with <25 donation-consistent deaths for a given race/ethnicity were excluded from analyses.

## Results

In 2019, OPO recovery rates varied substantially between the lowest- and highest-performing OPOs (Figure 1a), with greater variation for Asian (8.1-fold), non-Hispanic Black (NHB, 10.1-fold), and Hispanic (12.9-fold) populations than non-Hispanic whites (NHW, 3.7-fold; Supplementary Table 1).

**Figure 1a:**
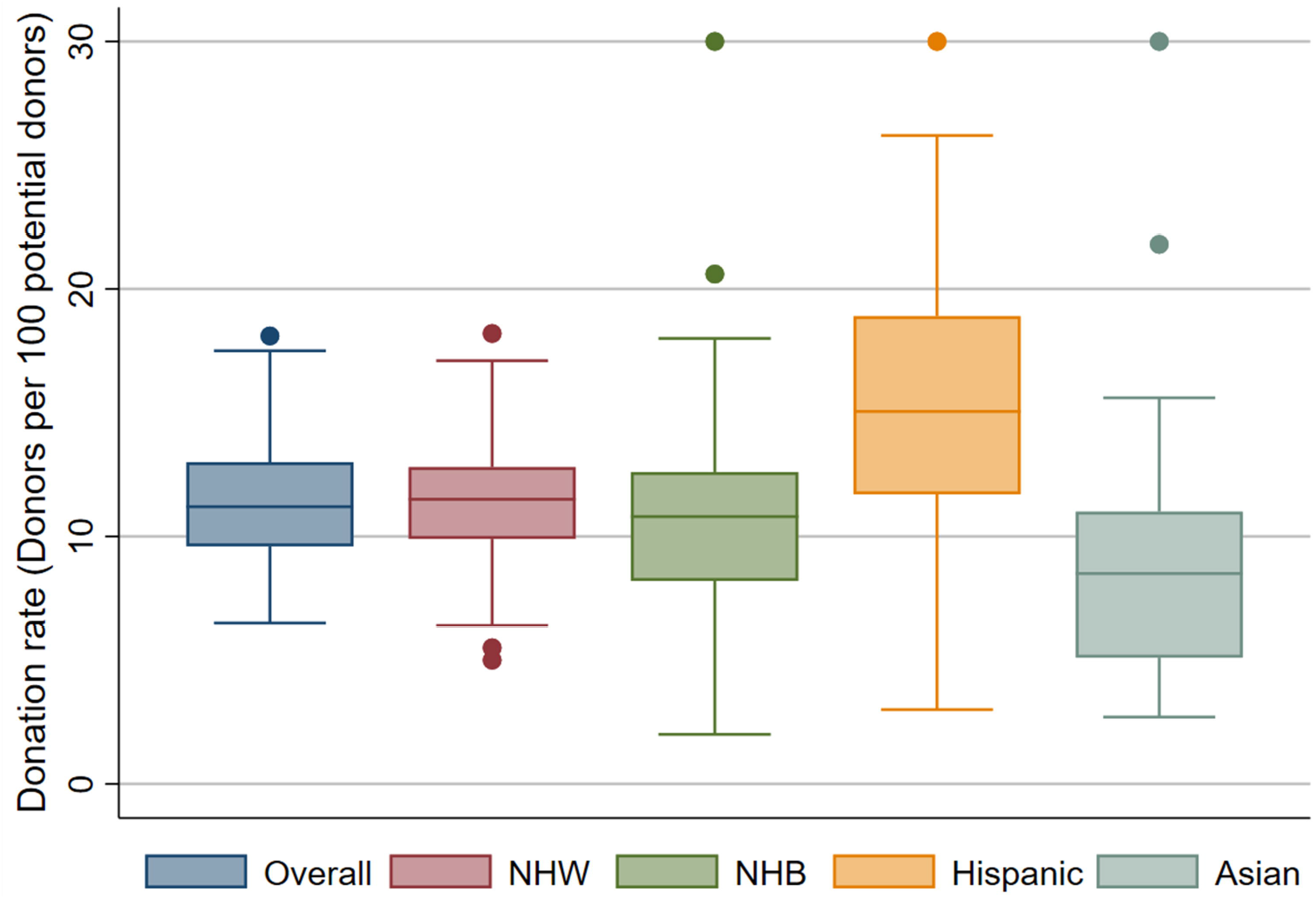
Box-and-whisker plots of OPO donation rate data (donors per 100 potential donors) in 2019 based on CMS OPO metrics.

**Figure 1b-1e:**
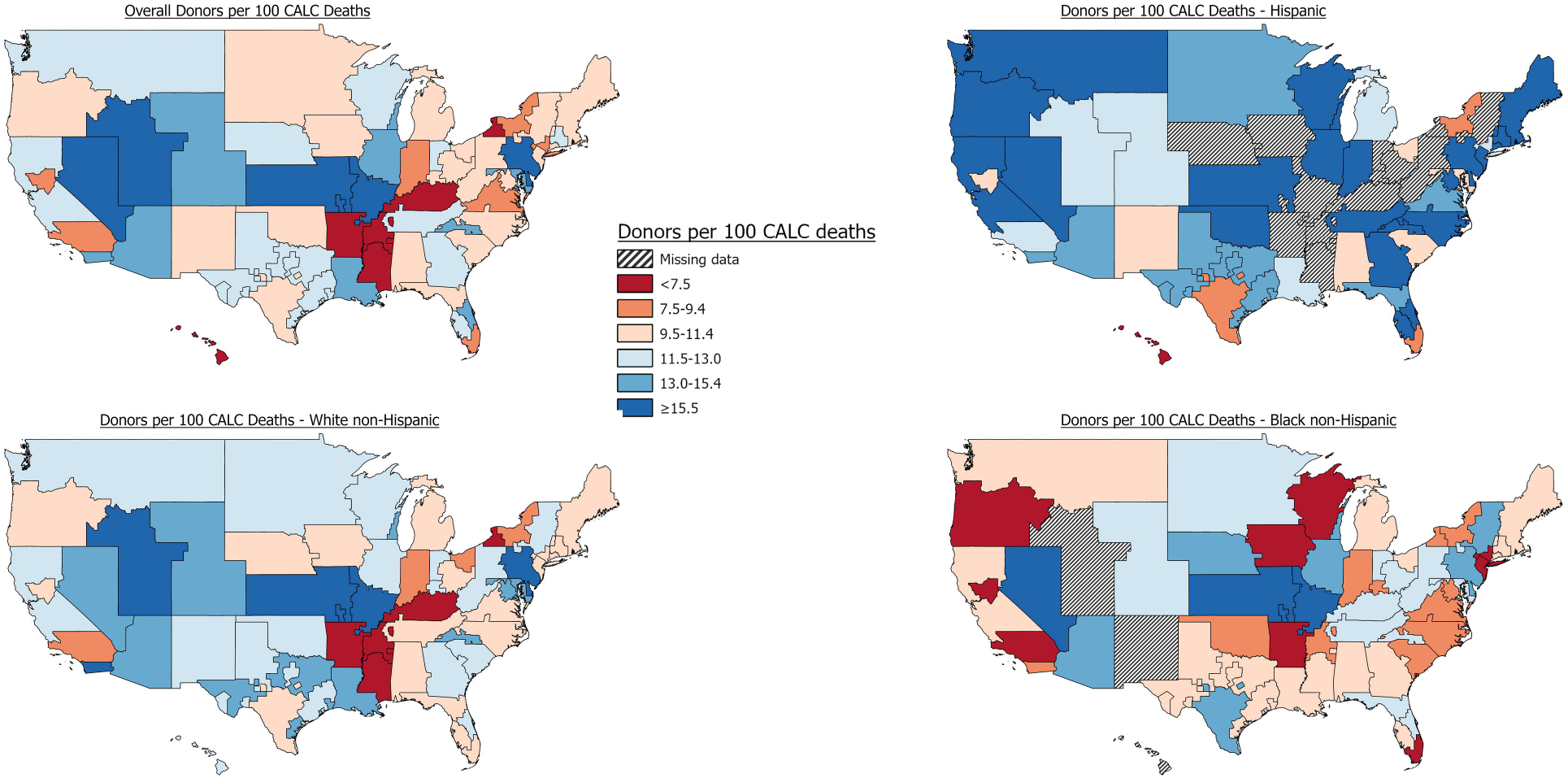
Organ procurement organization donation rate data (donors per 100 potential donors) in 2019 based on CMS OPO metrics*. *Donor service areas (DSAs) were excluded if there were <25 potential donors within a given race/ethnicity to avoid biased results from a small sample size. ‘Donation’ rates based on CMS metrics calculated as number of deceased donors (patient with ≥1 organ transplanted) per 100 potential donors (inpatient deaths ≤75 years of age who died from a cause consistent with donation). Box and whisker plots for NHW (n=l), NHB (n=l), and Asian (n=l) capped at 30 for graphical presentation. US map data not presented for Asian potential donors because only 28 OPOs had ≥25 potential Asian donors.

Of 16 Tier 3 OPOs per 2019 data, 15 are geographically adjacent to a Tier 1 OPO.^2^ For example, in Florida there are two Tier 3 OPOs: Life Alliance Organ Recovery Agency (Miami) and Life Quest Organ Recovery Services (Gainesville); and two Tier 1 OPOs: OurLegacy (Orlando) and LifeLink of Florida (Tampa). Though the two low-performing OPOs had similar NHW performance (20th percentile for Gainesville and 25^th^ percentile for Miami), performance with minorities was markedly different (75^th^ and 50^th^ percentiles, respectively for NHB and Hispanic populations in Gainesville DSA versus 5^th^ and 10^th^ percentile in Miami). Other low-performing OPOs with large minority populations had variable performance for NHW vs minority populations: We Are Sharing Hope (South Carolina, Tier 2) had higher donation rates for NHW (12.3 donors per 100 donation-consistent deaths; 65^th^ percentile) versus NHB (8.2 donors per 100 donation-consistent deaths; 25^th^ percentile), compared to OneLegacy (Los Angeles, Tier 3) where NHW white donation (8.5 donors per 100 donation-consistent deaths; 10^th^ percentile) lagged behind Hispanic donation (12.6 donors per 100 donation-consistent deaths; 35^th^ percentile).

## Discussion

Meeting new federal goals for increasing the organ supply will require tailored approaches to DSA-level variation in donor procurement. Race/ethnicity-stratified OPO-level data can identify populations among Tier 2 and 3 OPOs to focus quality improvement and outreach efforts in a process analogous to that used by a Tier 3 OPO to correct underperformance among older decedents.^5^ In Florida, for example, the interventions at the two Tier 3 OPOs should differ. In Gainesville, focus on NHW potential donors is needed, while the findings in Miami, where performance was low across all racial/ethnic groups, indicate process problems independent of decedent race/ethnicity.

Our study has limitations, including an inability to verify reported race/ethnicity and lack of process data to assess drivers of observed differences (e.g., variation in authorization approach across demographic groups). Nevertheless, the marked variability in race/ethnicity-specific procurement within and among OPOs contravenes assumptions that donation rates are lower in certain DSAs due to their demographics.^6^ These data can assist policymakers, OPOs, and clinicians in addressing deficits in donation and providing more organs for transplantation.

## Data Availability

All data produced in the present study are available upon reasonable request to the authors

## References

1. Medicare and Medicaid Programs; Organ Procurement Organizations Conditions for Coverage. https://www.federalregister.gov/documents/2021/02/02/2021-02180/medicare-and-medicaid-programs-organ-procurement-organizations-conditions-for-coverage-revisions-to. Accessed.

2. Center for Medicare and Medicare Services. S&C’s Quality, Certification and Oversight Reports (QCOR). Interim OPO Performance Report for the 2026 Certification Period, July 2021. https://qcor.cms.gov/main.jsp. Accessed October 6, 2021.

3. Doby BL, Brockmeier D, Lee KJ, et al. Opportunity to increase deceased donation for United States veterans. Am J Transplant. 2021.

4. Goldberg D, Karp S, Shah MB, Dubay D, Lynch R. Importance of incorporating standardized, verifiable, objective metrics of organ procurement organization performance into discussions about organ allocation. American Journal of Transplantation. 2019;19(11):2973–2978.

5. Doby BL, Hanner K, Johnson S, Purnell TS, Shah MB, Lynch RJ. Results of a data-driven performance improvement initiative in organ donation. American Journal of Transplantation. 2021;21(7):2555–2562.

6. Mone T, Danovitch G. US Organ Procurement Organization Donation Principles, Laws, and Practices. American journal of kidney diseases : the official journal of the National Kidney Foundation. 2020;76(5):735–738.

