## Supplemental Table 1 for "Identifying opportunities for improving the organ supply through race-stratified data"

Supplementary Table 1: Race/ethnicity stratified donation rates per OPO in 2019*

| OPO | Non-Hispanic white donation rate | Non-Hispanic Black donation rate | Hispanic donation rate | Asian donation rate |
| --- | --- | --- | --- | --- |
| ALOB | 10.6 | 9.7 | 11.1 | . |
| AROR | 6.4 | 6.6 | . | . |
| AZOB | 14.2 | 15.2 | 15.2 | 2.7 |
| CADN | 12.1 | 10.6 | 16.3 | 8.5 |
| CAGS | 10.1 | 6.2 | 9.8 | 4.8 |
| CAOP | 8.5 | 5.7 | 12.6 | 5.6 |
| CASD | 15.6 | 8.3 | 14.0 | 12.5 |
| CORS | 15.3 | 12.7 | 12.5 | 8.6 |
| CTOP | 11.0 | 9.8 | 20.2 | . |
| DCTC | 13.5 | 7.5 | 18.8 | 14.3 |
| FLFH | 12.4 | 12.1 | 21.2 | 10.8 |
| FLMP | 9.8 | 4.2 | 8.4 | 3.2 |
| FLUF | 9.6 | 12.6 | 14.9 | . |
| FLWC | 11.3 | 10.0 | 15.9 | 3.7 |
| GALL | 11.7 | 11.3 | 16.7 | 10.4 |
| HIOP | 12.4 | . | 3.0 | 6.2 |
| IAOP | 9.6 | 2.0 | . | . |
| ILIP | 12.8 | 13.6 | 17.5 | 10.4 |
| INOP | 8.9 | 8.2 | 16.0 | . |
| KYDA | 6.5 | 12.0 | . | . |
| LAOP | 15.0 | 10.9 | 11.9 | . |
| MAOB | 10.4 | 11.4 | 19.6 | 6.1 |
| MDPC | 14.7 | 11.9 | 10.8 | 7.7 |
| MIOP | 11.2 | 10.1 | 11.7 | 9.1 |
| MNOP | 11.8 | 12.9 | 14.7 | . |
| MOMA | 15.6 | 18.0 | . | 15.4 |
| MSOP | 5.5 | 9.7 | . | . |
| MWOB | 17.1 | 20.6 | 39.2 | . |
| NCCM | 14.3 | 11.6 | 24.2 | . |
| NCNC | 11.2 | 8.2 | 26.2 | 15.6 |
| NEOR | 11.0 | 13.3 | . | . |
| NJTO | 10.7 | 6.6 | 19.9 | 3.0 |
| NMOP | 12.0 | . | 10.1 | . |
| NVLV | 14.4 | 15.6 | 24.6 | 21.8 |
| NYAP | 11.8 | 13.8 | . | . |
| NYFL | 9.3 | 8.8 | 7.7 | . |
| NYRT | 9.9 | 7.2 | 12.1 | 7.1 |
| NYWN | 7.3 | . | 5.9 | . |
| OHLB | 9.3 | 10.8 | 10.7 | . |
| OHLC | 12.3 | 12.4 | . | . |
| OHLP | 10.1 | 12.5 | 20.0 | . |
| OHOV | 11.5 | 8.5 | . | . |
| OKOP | 12.1 | . | 16.7 | . |
| ORUO | 9.4 | 6.1 | 16.3 | 9.1 |
| PADV | 18.2 | 13.6 | 24.2 | 5.1 |
| PATF | 11.4 | 12.2 | . | . |
| SCOP | 12.3 | 8.2 | 10.6 | . |
| TNDS | 11.2 | 11.7 | 18.9 | . |
| TNMS | 5.0 | 8.0 | . | . |
| TXGC | 11.9 | 11.0 | 14.6 | 7.3 |
| TXSA | 10.3 | 14.4 | 8.6 | 3.8 |
| TXSB | 13.0 | 10.4 | 14.6 | 14.9 |
| UTOP | 16.1 | . | 12.2 | . |
| VATB | 9.5 | 8.9 | 14.7 | 3.4 |
| WALC | 12.5 | 10.5 | 17.3 | 11.0 |
| WIDN | 14.0 | 14.6 | 24.6 | . |
| WIUW | 12.7 | 6.5 | 16.0 | . |

* Donation rate calculated using CMS methodology: deceased donors (patient with ≥1 organ transplanted) / donation consistent deaths (inpatient deaths ≤75 years of age from causes consistent with death using National Center for Health Statistics data). To avoid small sample size bias, DSAs with <25 donation-consistent deaths for a given race/ethnicity were excluded from analyses.
